# Genetic Susceptibility as an Effect Modifier for the Association of Chronic Kidney Disease with Risks of Venous Thromboembolism and Pulmonary Embolism

**DOI:** 10.1101/2023.02.27.23286539

**Authors:** Jingwen Zhang, Ying Shan, Beini Liu, Liang Dai, Simin Du, Congying Song, Jianqing Shi, Juan Jesus Carrero, Zuying Xiong, Xiaoyan Huang

## Abstract

**Background:** The association between chronic kidney disease (CKD) and the risk of developing venous thromboembolism (VTE) or pulmonary embolism (PE) is still controversial. Further, it is unknown if genetic predisposition modifies these relationships. This work aimed to investigate plausible effect modification of genetic factors on the association between CKD and incident VTE, as well as incident PE.

**Methods:** Population-based cohort study of the UK Biobank, including participants of European ancestry that were free of VTE (397,658 participants) or PE (406,486 participants) at recruitment. We used cystatin C combining creatinine estimated glomerular filtration rate and albuminuria to classify the participants as the low, intermediate, and high or very high-risk groups suggested by KDIGO. Cox proportional hazards model was applied to evaluate the associations of CKD with incident VTE and PE. In addition, we used an externally validated polygenic risk score (PRS) for VTE to evaluate whether the genetic predisposition modified the associations of interest.

**Results:** During median follow-up of 12.7 years, 11,372 participants developed VTE, and 6,518 participants developed PE. As compared with the low KDIGO risk category, covariate-adjusted hazard ratios (HR) and 95% confidence intervals (CI) for VTE risk with the intermediate KDIGO risk category and high or very high KDIGO risk category at baseline were 1.278 (1.191-1.372) and 1.892 (1.658-2.159), respectively. Participants at high or very high KDIGO risk category and in the highest tertile of PRS had the highest risk of developing VTE (HR, 4.397; 95% CI, 3.639-5.313), and this group showed the most conspicuous additive interaction between CKD and the genetic predisposition, which was responsible for 1.389-fold relative excess risk and 31.6% of the VTE risk. Also, when either estimated glomerular filtration rate or urine albumin-creatinine ratio was treated as the exposure, consistent associations were observed. Analyses for PE yielded similar associations and supra-additive interactions.

**Conclusions:** CKD is associated with future VTE and PE, especially in those with high genetic risk.

**CLINICAL PERSPECTIVE:** 

**What is new?:**

- We observed that chronic kidney disease is associated with future venous thromboembolism and pulmonary embolism independently, irrespective of genetic predisposition.
- We found considerable additive interaction between chronic kidney disease and genetic predisposition for future venous thromboembolism or pulmonary embolism risk.

**What are the clinical implications?:**

- It is essential to evaluate kidney condition in primary prevention of venous thromboembolism and pulmonary embolism.
- Individuals with high genetic risk of venous thromboembolism might be the most relevant population to implement a personalized program of chronic kidney disease prevention, screening, and management.

## INTRODUCTION

Venous thromboembolism (VTE), afflicting nearly 10 million people worldwide per year, is a significant contributor to mortality.^1^ The estimated annual health-care costs related with VTE range between 1.5-3.3 billion EUR in Europe^2^ and 7-10 billion USD in the United States.^3^ VTE comprises both deep vein thrombosis and pulmonary embolism (PE), the latter of which is responsible for most VTE - related deaths. Only approximately 20% of all VTE are triggered by strong provoking factors like active cancer or prolonged immobilization, whereas the remaining VTE events are provoked by multiple weak risk factors or are unprovoked (no apparent risk factor).^4,5^ Thus, unraveling more common risk factors would be essential for reducing the global burden of VTE.

VTE is believed to be a multifactorial disease, with genetic and acquired risk factors as well as possibly their interplay contributing to the development of VTE. Individuals with chronic kidney disease (CKD) commonly manifest a prothrombotic state, indicating that kidney impairment may be an emerging risk factor for VTE.^6^ CKD, defined as estimated glomerular filtration rate (eGFR) < 60 ml/min/1.73 m^2^ or urine albumin-creatinine ratio (UACR) ≥ 30 mg/g, is reported to associate with high risk of VTE in a pooled analysis of 5 community-based cohorts.^7^ Of note, low eGFR^7,8^ and high UACR^7,9^ even within their normal ranges are independently associated with incident VTE in the general population. What is more, a number of susceptibility loci for VTE have been revealed by recent genome-wide association studies (GWAS).^10,11^

Most of these loci are located in or near genes encoding coagulation proteins. Accordingly, polygenic risk scores (PRS) combining single nucleotide polymorphisms (SNP) have been constructed and allow personalized measure of genetic liability of VTE. However, it remains unknown if kidney disorders aggravate the genetic risk of VTE.

We hypothesized that genetic predisposition modifies the relationship between CKD and VTE or PE. In this prospective cohort study of community-dwelling participants, we investigated the associations of CKD according to the KDIGO risk category of CKD prognosis and its two key markers, ie, eGFR and UACR, with the incidence of both VTE and PE. Furthermore, we quantitated both additive and multiplicative interactions to assess effect modification by VTE-related PRS.

## METHODS

### Study design

This is a prospective cohort study based on UK Biobank. The UK Biobank is a national longitudinal cohort study, which recruited more than 500,000 individuals aged between 40 and 69 years from 22 assessment centers in the UK general population during 2006 - 2010.^12^ The UK Biobank obtained ethics approval from the North West Multi-Centre Research Ethics Committee (11/NW/0382), and informed consent was obtained from all participants. Baseline information was gathered through touchscreens, nurse-led questionnaires, physical measurements and biological samples. Future health status including disease incidence, hospitalization, and death was also monitored.

### Study population

For this study, we included the UK Biobank participants who were Whites of European ancestry that were free of VTE or PE at baseline and did not withdraw until November 12^th^ 2021. Participants who met any of the following criteria were excluded: (1) did not have data on PRS for VTE; (2) with missing data to calculate eGFR or UACR, such as age, sex, serum cystatin C, serum creatinine, urinary albumin, or urinary creatinine; (3) with prevalent end-stage kidney disease at baseline; (4) hospitalized and diagnosed with acute kidney failure within 90 days around baseline. The process for the analytical cohort construction is shown in the **Figure S1**.

### Kidney measures

Serum and random spot urine samples were collected at baseline and analyzed at a central laboratory. A detailed description of the assay methods and quality assurance protocols can be found online (https://biobank.ndph.ox.ac.uk/showcase/showcase/docs/serum_biochemistry.pdf). In the main analysis, the participants were grouped, according to both eGFR and UACR levels at baseline, into low risk, intermediate risk, high or very high risk in line with prognosis risk stratification recommended by the 2012 KDIGO (Kidney Disease: Improving Global Outcomes) guideline.^13^ Due to the limited samples with KDIGO high and very high risk categories in the UK Biobank, we determined a priori to pool them in this study. eGFR or UACR along was treated as secondary exposure variables. eGFR was calculated using the Chronic Kidney Disease Epidemiology Collaboration (CKD-EPI) eGFR_cr-cys_ refit equation which combined both serum creatinine and cystatin C without race variable^14^, and categorized as <60, 60 -<90, ≥90 ml/min/1.73 m^2^. UACR was categorized as <30, 30-300, >300 mg/g.

### Polygenic risk score for venous thromboembolism

We used the standard PRS set for VTE released by the UK Biobank. In brief, the standard PRS set was calculated for all UK Biobank individuals and trained on external data. By constructing a standardized testing subgroup within the UK Biobank and a standardized set of disease and quantitative trait definitions, the PRSs were evaluated under a unified pipeline.^15^ In this study, the PRS for VTE was categorized based on the tertiles of the PRS among the non-VTE participants as low, intermediate, and high genetic risk.

### Outcome ascertainment

The disease outcomes were defined using International Classification of Diseases 10^th^ edition (ICD-10) codes, collected through inpatient hospital, primary care, self-report, and death registry data linked to the UK Biobank. The primary outcome was first reported fatal or nonfatal VTE (ICD-10: I26, I80-I82). The secondary outcome was first reported fatal or nonfatal PE (ICD-10: I26) **(Table S1)**.^16^ As per the 2019 European Society of Cardiology/European Respiratory Society guidelines,^17^ we avoided separating “provoked” and “unprovoked” VTE in the present study. The follow-up time was calculated from the date of recruitment until the date of incident VTE or PE, death, loss to follow-up, or November 12^th^ 2021, whichever occurred first.

### Statistical analysis

Baseline characteristics of the included participants are presented as mean (standard deviation) for normally distributed variables, median (interquartile range) for non-normally distributed variables, and frequency (percentage) for categorical variables. The incidence rates per 100,000 person-years were calculated. Multiple imputation was applied to impute the missing data of covariates according to different variable types. (predictive mean matching method for continuous variables imputation; logistic regression method for binary variables imputation; polytomous regression for polytomous categorical variables imputation). The number of imputations replications was 10. Univariate and covariate-adjusted Cox proportional hazards models were applied to analyze the association between CKD measures and the incidence of VTE or PE. Lost to follow-up was treated as a censoring event. The hazard ratios (HR) and 95% confidence intervals (95% CI) were reported. In the full-adjusted models, confounders we considered on the basis of prior literature included age, sex, highest attained education (higher, lower), Townsend deprivation index, smoking (never, previous, current), alcohol consumption (never, occasional, frequent), metabolic equivalents of physical activity, body mass index, hypertension (yes, no), diabetes (yes, no), cardiovascular diseases other than hypertension (yes, no), cancer (yes, no), cholesterol-lowering medications (yes, no), anticoagulants (yes, no), and antiplatelet agents (yes, no). In models for eGFR or UACR, we further mutually adjusted for each other. Linear trend analyses were also applied to detect the risk of VTE or PE risks across the KDIGO risk categories, and also across categories of eGFR or UACR. Variance inflation factors were used to measure multicollinearity. The proportional hazards assumption was assessed by visualizing the Schoenfeld residual.

To evaluate potential effect modification, we stratified the individuals into low, intermediate, and high genetic risk categories by tertile of PRS for standard VTE, and the models were further adjusted for the top 20 genetic principal components and genotyping batch. Multiplicative interaction was tested by including interaction terms between CKD and VTE-related PRS. We also assessed additive interaction by calculating the relative excess risk due to interaction (RERI) and the attributable proportion (AP).^18^ The 95% CI of RERI and AP were generated by drawing 1,000 bootstrap samples from the estimation data set.^19^

Sensitivity analyses were conducted to test the robustness of our results: (1) excluding participants with missing covariates; (2) excluding participants who developed VTE within the first year of follow-up; (3) excluding participants taking anticoagulants or antiplatelet agents at baseline; and (4) confirming all the analyses with the Fine and Gray models to handle the competing risk from pre-VTE deaths.

All analyses were performed with R version 4.0.3. All *P* values were two tailed, and a *P* value <0.05 was considered statistically significant.

## RESULTS

Characteristics of the study population for the primary outcome VTE analysis are shown in **Table 1**. A total of 397,658 participants were included, comprising 47.19% males. Mean age at baseline was 56.68 (standard deviation, 8.04) years, and most participants had relatively healthy kidney condition (94.32% participants with low KDIGO risk). Compared with participants with low KDIGO risk, those with high or very high KDIGO risk at baseline tended to be older and more obese, more likely to be current smokers but less likely to be current drinkers. They also presented to have more comorbidities and more likely to be on antiplatelet or anticoagulant drugs. During follow-up (median 12.7 years, 4,904,202 person-years at risk), a total of 11,372 (2.86%) participants developed VTE. For PE analysis, a total of 406,486 participants were included, comprising 47.01% males. Mean age at baseline was 56.74 (standard deviation, 8.03) years, and 94.32% participants with low KDIGO risk). During follow-up (median 12.73 years, 5,036,673 person-years at risk), 6,518 (1.60%) participants developed PE.

**Table 1.**
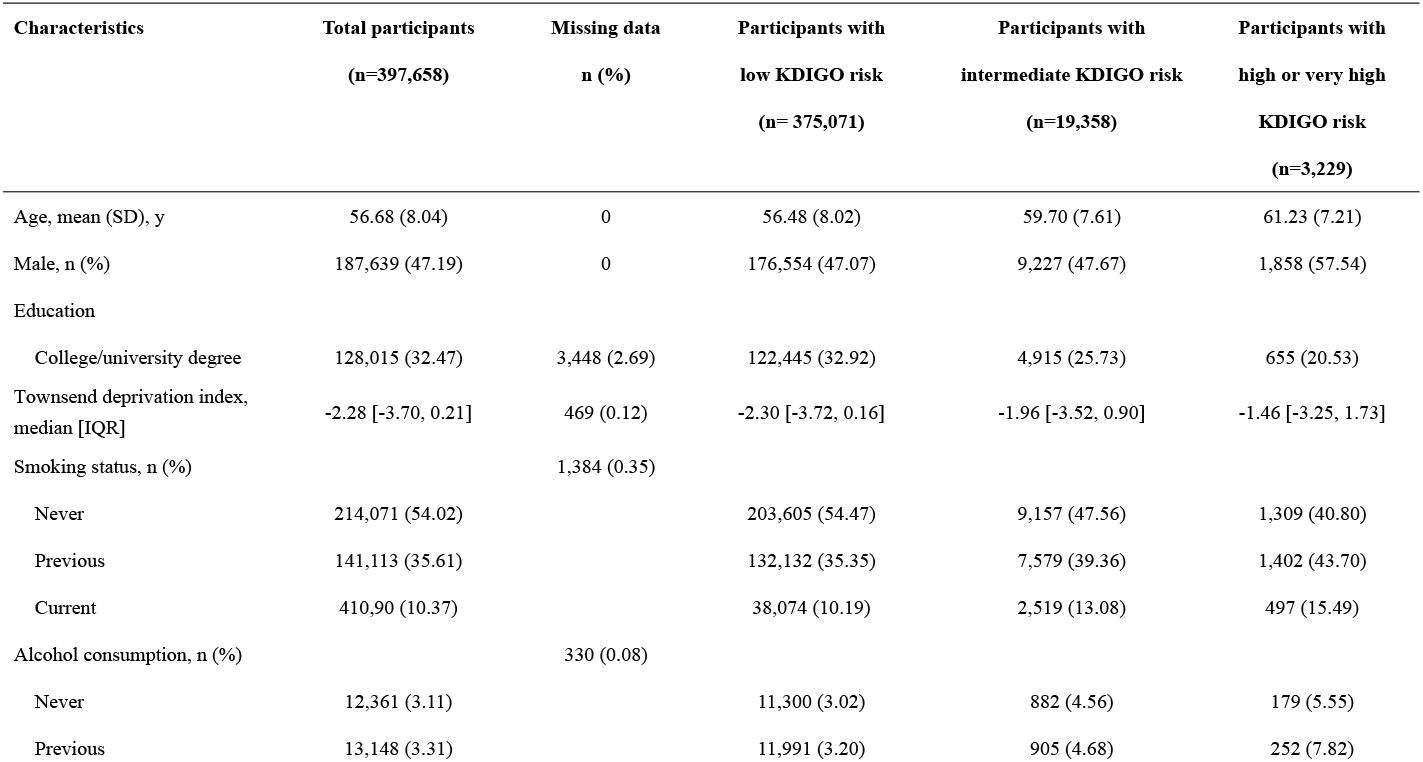
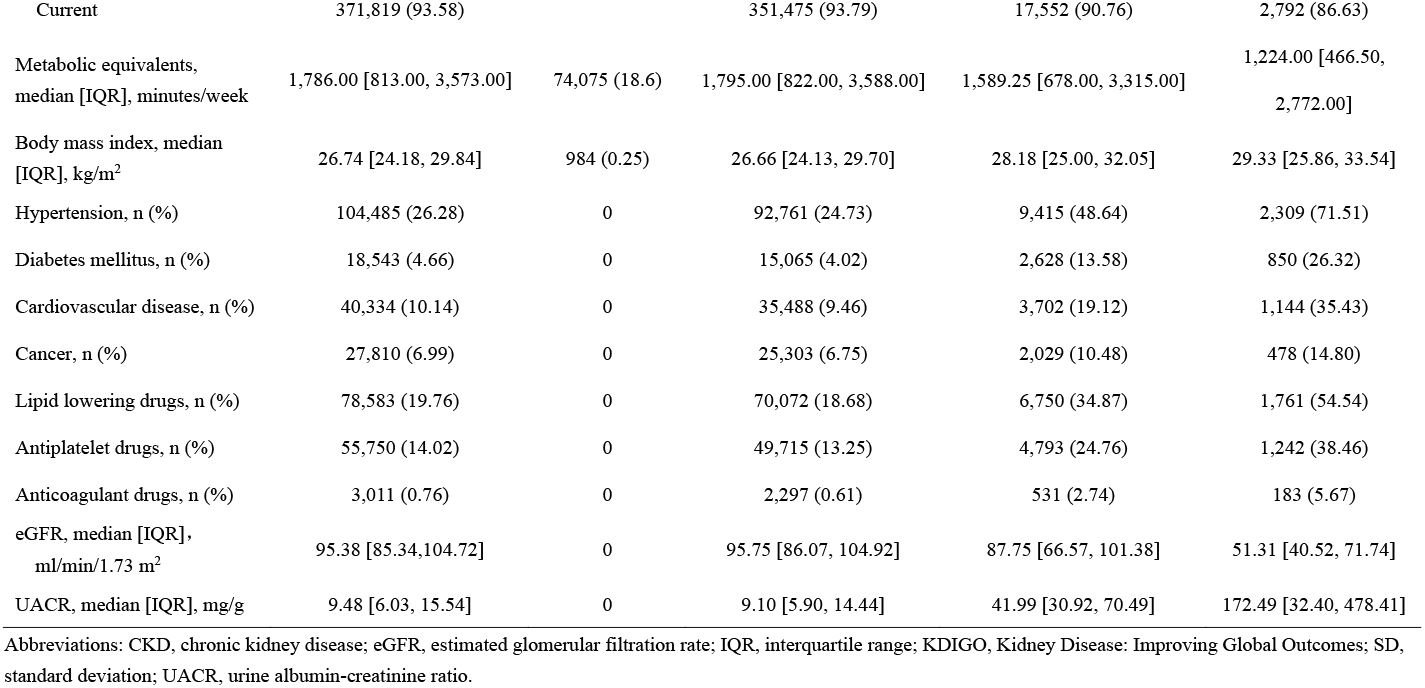
Baseline characteristics by KDIGO risk categories for venous thromboembolism risk analysis

| **Characteristics** | **Total participants**<br>**(n=397,658)** | **Missing data**<br>**n (%)** | **Participants with**<br>**low KDIGO risk**<br>**(n= 375,071)** | **Participants with**<br>**intermediate KDIGO risk**<br>**(n=19,358)** | **Participants with**<br>**high or very high**<br>**KDIGO risk**<br>**(n=3,229)** |
| --- | --- | --- | --- | --- | --- |
| Age, mean (SD), y | 56.68 (8.04) | 0 | 56.48 (8.02) | 59.70 (7.61) | 61.23 (7.21) |
| Male, n (%) | 187,639 (47.19) | 0 | 176,554 (47.07) | 9,227 (47.67) | 1,858 (57.54) |
| Education |  |  |  |  |  |
| College/university degree | 128,015 (32.47) | 3,448 (2.69) | 122,445 (32.92) | 4,915 (25.73) | 655 (20.53) |
| Townsend deprivation index,<br>median [IQR] | -2.28 [-3.70, 0.21] | 469 (0.12) | -2.30 [-3.72, 0.16] | -1.96 [-3.52, 0.90] | -1.46 [-3.25, 1.73] |
| Smoking status, n (%) |  | 1,384 (0.35) |  |  |  |
| Never | 214,071 (54.02) |  | 203,605 (54.47) | 9,157 (47.56) | 1,309 (40.80) |
| Previous | 141,113 (35.61) |  | 132,132 (35.35) | 7,579 (39.36) | 1,402 (43.70) |
| Current | 410,90 (10.37) |  | 38,074 (10.19) | 2,519 (13.08) | 497 (15.49) |
| Alcohol consumption, n (%) |  | 330 (0.08) |  |  |  |
| Never | 12,361 (3.11) |  | 11,300 (3.02) | 882 (4.56) | 179 (5.55) |
| Previous | 13,148 (3.31) |  | 11,991 (3.20) | 905 (4.68) | 252 (7.82) |
| Current | 371,819 (93.58) |  | 351,475 (93.79) | 17,552 (90.76) | 2,792 (86.63) |
| Metabolic equivalents,<br>median [IQR], minutes/week | 1,786.00 [813.00, 3,573.00] | 74,075 (18.6) | 1,795.00 [822.00, 3,588.00] | 1,589.25 [678.00, 3,315.00] | 1,224.00 [466.50,<br>2,772.00] |
| Body mass index, median<br>[IQR], kg/m<sup>2</sup> | 26.74 [24.18, 29.84] | 984 (0.25) | 26.66 [24.13, 29.70] | 28.18 [25.00, 32.05] | 29.33 [25.86, 33.54] |
| Hypertension, n (%) | 104,485 (26.28) | 0 | 92,761 (24.73) | 9,415 (48.64) | 2,309 (71.51) |
| Diabetes mellitus, n (%) | 18,543 (4.66) | 0 | 15,065 (4.02) | 2,628 (13.58) | 850 (26.32) |
| Cardiovascular disease, n (%) | 40,334 (10.14) | 0 | 35,488 (9.46) | 3,702 (19.12) | 1,144 (35.43) |
| Cancer, n (%) | 27,810 (6.99) | 0 | 25,303 (6.75) | 2,029 (10.48) | 478 (14.80) |
| Lipid lowering drugs, n (%) | 78,583 (19.76) | 0 | 70,072 (18.68) | 6,750 (34.87) | 1,761 (54.54) |
| Antiplatelet drugs, n (%) | 55,750 (14.02) | 0 | 49,715 (13.25) | 4,793 (24.76) | 1,242 (38.46) |
| Anticoagulant drugs, n (%) | 3,011 (0.76) | 0 | 2,297 (0.61) | 531 (2.74) | 183 (5.67) |
| eGFR, median [IQR],<br>ml/min/1.73 m<sup>2</sup> | 95.38 [85.34, 104.72] | 0 | 95.75 [86.07, 104.92] | 87.75 [66.57, 101.38] | 51.31 [40.52, 71.74] |
| UACR, median [IQR], mg/g | 9.48 [6.03, 15.54] | 0 | 9.10 [5.90, 14.44] | 41.99 [30.92, 70.49] | 172.49 [32.40, 478.41] |

Abbreviations: CKD, chronic kidney disease; eGFR, estimated glomerular filtration rate; IQR, interquartile range; KDIGO, Kidney Disease: Improving Global Outcomes; SD, standard deviation; UACR, urine albumin-creatinine ratio.

### Risk associations between kidney measures and study outcomes

As the “KDIGO risk heat map” (**Figure 1**) depicts, the incidence rates of VTE and PE became progressively higher with a worse stages of CKD, lower eGFR as well as higher UACR categories. **Table 2** presents the associations of KDIGO risk categories with risk of VTE and PE. For VTE risk, as compared with the group with low KDIGO risk, fully-adjusted HR and 95% CI for participants with intermediate and high or very high KDIGO risk were 1.278 (1.191-1.372) and 1.892 (1.658-2.159), respectively. These associations were robust after excluding participants with missing covariates, participants developing VTE within the first year of follow-up, or participants taking anticoagulants or antiplatelet agents at baseline **(Table S2)**. We also considered the possible competing risk from non-VTE death using Fine and Gray models, the associations were slightly attenuated **(Table S2)**. Both eGFR and UACR were also associated with incident VTE or PE independently **(Table S3)**.

**Table 2.**
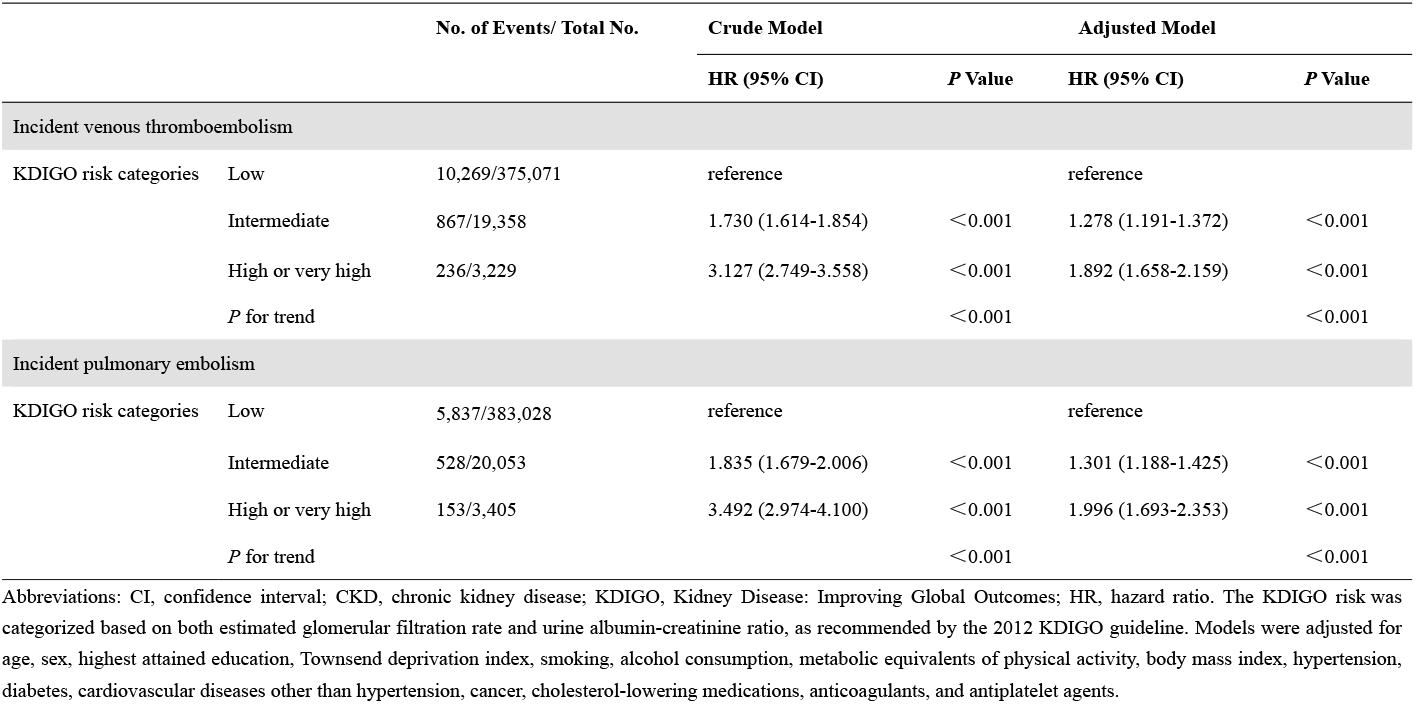
Associations between KDIGO risk categories and risks of venous thromboembolism and pulmonary embolism

|  |  |  | Crude Model |  | Adjusted Model |  |
| --- | --- | --- | --- | --- | --- | --- |
|  |  |  | HR (95% CI) | P Value | HR (95% CI) | P Value |
| Incident venous thromboembolism |  |  |  |  |  |  |
| KDIGO risk categories | Low | 10,269/375,071 | reference |  | reference |  |
|  | Intermediate | 867/19,358 | 1.730 (1.614-1.854) | <0.001 | 1.278 (1.191-1.372) | <0.001 |
|  | High or very high | 236/3,229 | 3.127 (2.749-3.558) | <0.001 | 1.892 (1.658-2.159) | <0.001 |
|  | *P* for trend |  |  | <0.001 |  | <0.001 |
| Incident pulmonary embolism |  |  |  |  |  |  |
| KDIGO risk categories | Low | 5,837/383,028 | reference |  | reference |  |
|  | Intermediate | 528/20,053 | 1.835 (1.679-2.006) | <0.001 | 1.301 (1.188-1.425) | <0.001 |
|  | High or very high | 153/3,405 | 3.492 (2.974-4.100) | <0.001 | 1.996 (1.693-2.353) | <0.001 |
|  | *P* for trend |  |  | <0.001 |  | <0.001 |

Abbreviations: CI, confidence interval; CKD, chronic kidney disease; KDIGO, Kidney Disease: Improving Global Outcomes; HR, hazard ratio. The KDIGO risk was categorized based on both estimated glomerular filtration rate and urine albumin-creatinine ratio, as recommended by the 2012 KDIGO guideline. Models were adjusted for age, sex, highest attained education, Townsend deprivation index, smoking, alcohol consumption, metabolic equivalents of physical activity, body mass index, hypertension, diabetes, cardiovascular diseases other than hypertension, cancer, cholesterol-lowering medications, anticoagulants, and antiplatelet agents.

**Figure 1.**
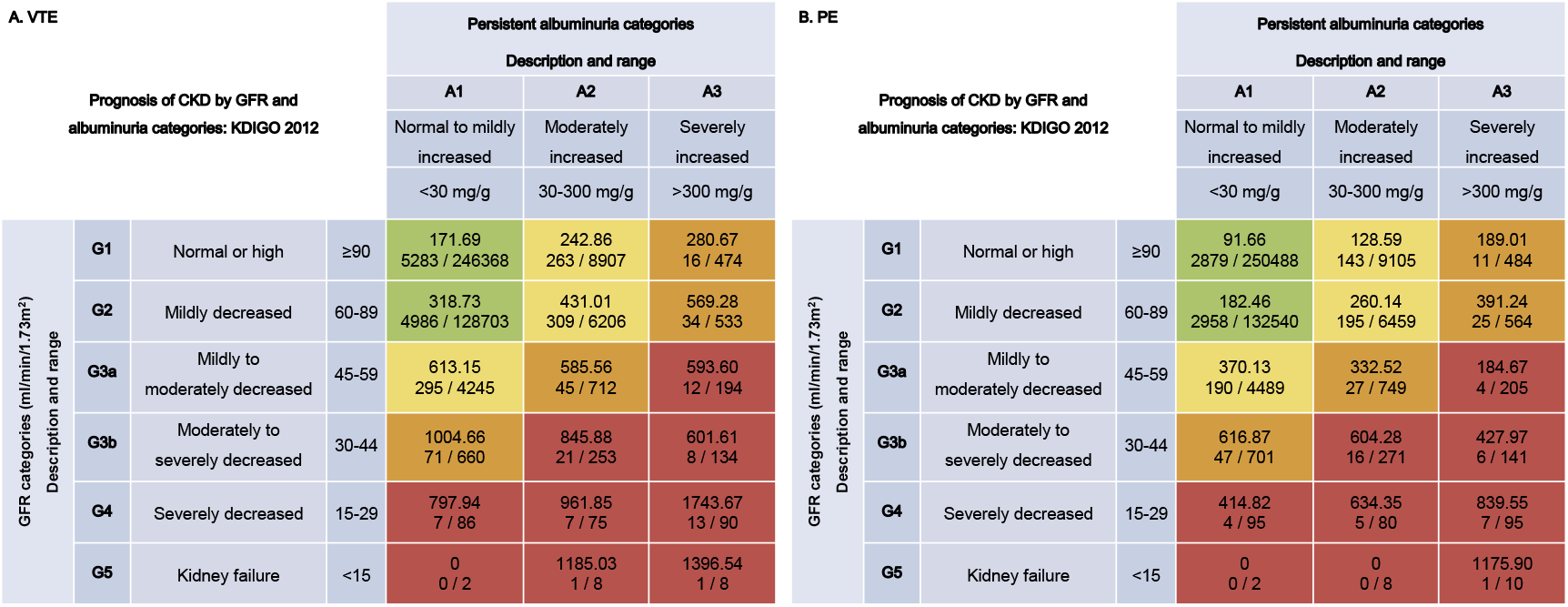
Distribution and incidence of venous thromboembolism and pulmonary embolism events. Abbreviations: CKD, chronic kidney disease; GFR, glomerular filtration rate; KDIGO, Kidney Disease: Improving Global Outcomes; PE, pulmonary embolism; VTE, venous thromboembolism. The participants were grouped according to KDIGO risk categories based on both estimated glomerular filtration rate and urine albumin-creatinine ratio, as recommended by the 2012 KDIGO guideline. Green: low risk (if no other markers of kidney disease, no chronic kidney disease); Yellow: moderately increased risk; Orange: high risk; Red, very high risk. Numbers in each cell are incidence rate (per 100,000 person-year) and events / sample size of venous thromboembolism (Panel A) and pulmonary embolism (Panel B).

### Risk associations between VTE-related polygenic risks scores and study outcomes

VTE-related PRS of participants ranges from -3.98 to 7.16, participants were grouped with PRS tertiles (−0.44 and 0.39) into low, intermediate and high genetic risk. Participants in the high genetic risk susceptibility (top tertile) category had a significantly higher risk of VTE (HR, 2.184; 95% CI, 2.083–2.290) and PE (HR, 2.508; 95% CI, 2.351–2.675) as compared with those in the low genetic risk category **(Table S4)**. When stratifying the participants by their genetic predisposition, we observed that the associations of CKD, as well as eGFR and UACR, with VTE or PE risk increased in a dose-response manner. The VTE or PE risk was incrementally increased across worsening higher genetic risk categories and worse CKD categories (either KDIGO risk groups, lower eGFR, or higher UACR categories) (**Figure 2, Figure S2**), and these patterns were consistent in all sensitivity analyses **(Table S5-S8)**. Specifically, compared with participants with low KDIGO risk category and in the lowest tertile of PRS, those at high or very high KDIGO risk category and concurrently in the highest tertile of PRS had the highest risk of VTE (HR, 4.397; 95% CI, 3.639-5.313) and PE (HR, 5.093; 95% CI, 4.015 - 6.459) (**Figure 2**).

**Figure 2.**
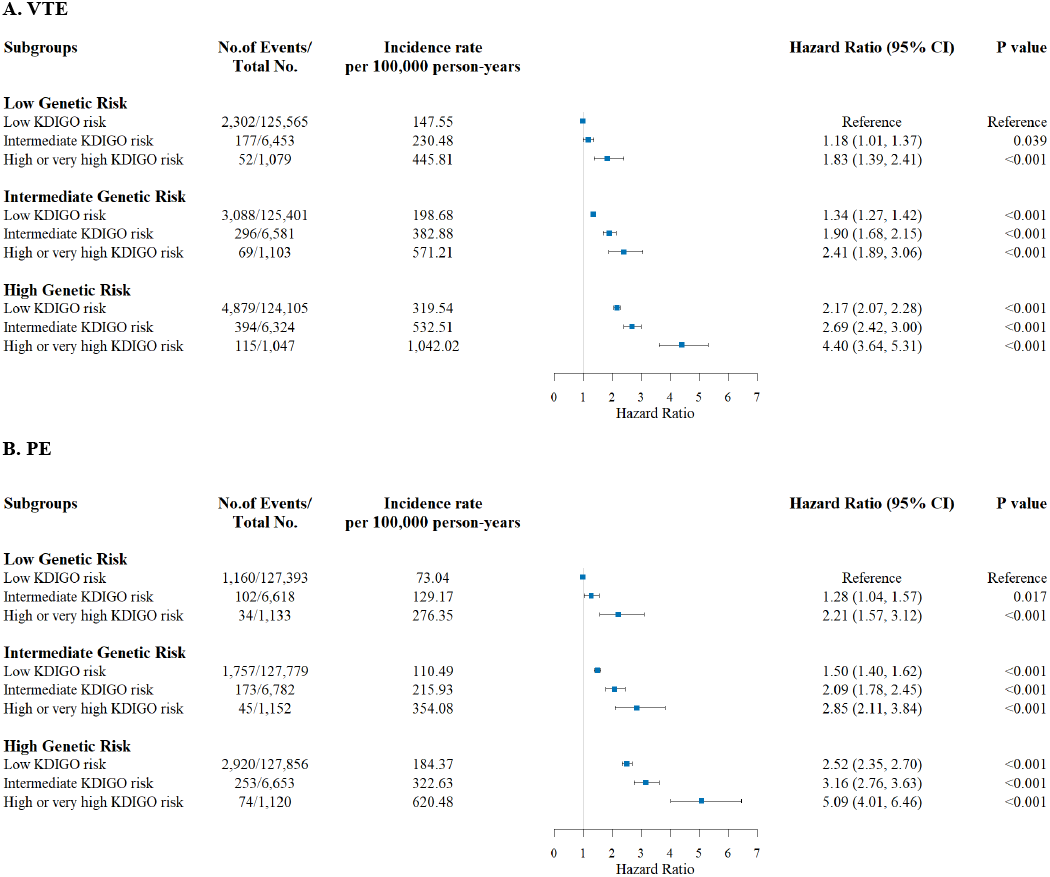
Venous thromboembolism and pulmonary embolism risks according to genetic susceptibility and KDIGO risk categories. Abbreviations: CI, confidence interval; CKD, chronic kidney disease; KDIGO, Kidney Disease: Improving Global Outcomes; PE, pulmonary embolism; VTE, venous thromboembolism. Hazard ratios were obtained from Cox proportional hazards models, and all models were adjusted for age, sex, highest attained education, Townsend deprivation index, smoking, alcohol consumption, metabolic equivalents of physical activity, body mass index, hypertension, diabetes, cardiovascular diseases other than hypertension, cancer, cholesterol-lowering medications, anticoagulants, antiplatelet agents, top 20 principal components of ancestry, and genotyping batch.

### Interactions between CKD severity and VTE-related polygenic risk score

We observed supra-additive interactions of CKD with VTE-related PRS, evident by positive RERI and AP values. Specifically, compared to low KDIGO risk category participants in the lowest tertile of PRS, RERI and AP for those at high or very high KDIGO risk category and concurrently in the highest tertile of PRS was 1.389 (95% CI, 0.439-2.346) and 0.316 (95% CI, 0.124-0.478), respectively (**Panel A of Figure 3, Panel A and C of Figure S3**). These estimates suggested that the interaction was responsible for 1.389-fold relative excess risk and accounted for 31.6% of the VTE risk. Analyses for PE yielded similar supra-additive interactions (**Panel B of Figure 3, Panel B and D of Figure S3**). We did not observe multiplicative interaction between PRS and the CKD measures **(Table S9)**. Sensitive analyses showed that the results were marginally changed **(Table S10-S11)**.

**Figure 3.**
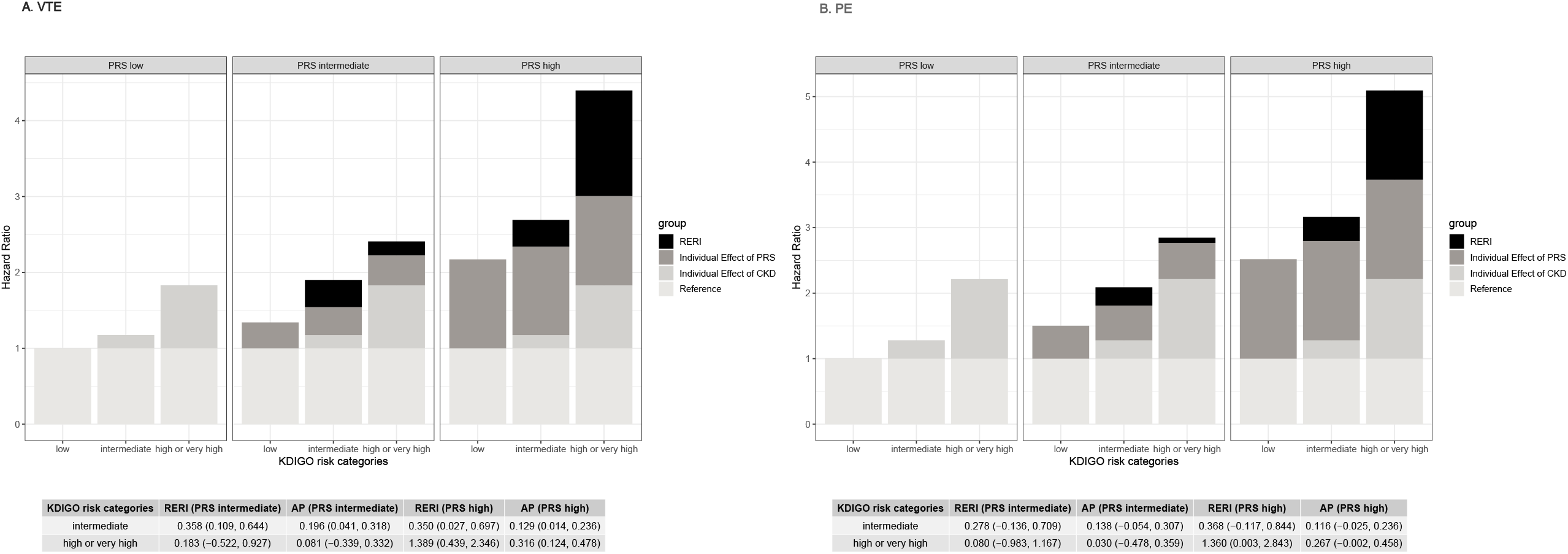
Additive interaction between KDIGO risk categories and genetic susceptibility on venous thromboembolism and pulmonary embolism risks. Abbreviations: AP, attributable proportion; CKD, chronic kidney disease; KDIGO, Kidney Disease: Improving Global Outcomes; PE, pulmonary embolism; PRS, polygenic risk score; RERI, relative excess risk due to interaction; VTE, venous thromboembolism. Reference was the group of low polygenic risk score and low risk of chronic kidney disease prognosis. All models were adjusted for age, sex, highest attained education, Townsend deprivation index, smoking, alcohol consumption, metabolic equivalents of physical activity, body mass index, hypertension, diabetes, cardiovascular diseases other than hypertension, cancer, cholesterol-lowering medications, anticoagulants, antiplatelet agents, top 20 principal components of ancestry, and genotyping batch.

## DISCUSSION

### Principal findings

In this prospective cohort study of community-dwelling participants from the UK Biobank, we witnessed dose-response relationships of KDIGO risk categories with incident VTE in 397,658 participants and incident PE in 406,486 participants. Either of suboptimal eGFR or UACR may contribute to such associations. The associations of the CKD with VTE or PE existed irrespective of genetic predisposition; however, the interaction between CKD and the genetic predisposition appeared considerable and supra-additive, ie, RERI reached as high as 1.389 and accounted for 31.6% of the observed VTE risk.

### Comparison with prior literature

Our results using the KDIGO recommended CKD prognosis stratification nicely complement existing evidence of VTE and, further, extend to the link with PE. Several observational studies have shown that low eGFR is independently associated with future VTE risk.^7,8,20-24^ Moreover, a recent Mendelian randomization study indicates this association may be causal.^25^ Our results also support the association between albuminuria and risk of VTE, which are consistent with most,^7,9,24^ if not all,^8,21^ of prior studies. A meta-analysis conducted by Mahmoodi et al.^7^ in 2012 pooled individual participant data from 5 community-based cohorts from Europe and the United States (n= 95,154 participants included in total). The authors concluded both eGFR and UACR even within the normal ranges are independently associated with increased VTE risk in the general population.

Our analysis on gene-kidney interaction provides novel insights into the etiology of VTE. It is widely accepted that VTE is affected by both inherited and environmental factors. The heritability of VTE is estimated to range between 23%^10^ and 30%^11^. Common genetic variants selected from GWAS, including those in the *ABO, F2, F5, F11, FGG*, or *ZFPM2* genes, may affect the blood coagulability and hemostasis traits.^26-30^ In addition, the PRS that combined multiple SNP showed a dose-response association with the population-level risk of VTE in the present study. Although previous research suggests that VTE risk is greatest when genetic predisposition is combined with an environmental risk factor, little attention has been paid to date to the gene-kidney interaction. We observed a noteworthy supra-additive interaction between the PRS and kidney related traits, indicating a synergetic effect of these factors on VTE risk.

How the kidney interacts to influence VTE risk can be complex and is yet to be fully elucidated. In individuals with kidney diseases, mixed hemostatic abnormalities such as activation of procoagulants, decreased endogenous anticoagulants, platelet dysfunction, and reduced fibrinolytic activity have been reported.^31^ Elevated levels of procoagulant and proinflammatory substances, eg, D-dimer, factor VII, factor VIII, von Willebrand factor, C-reactive protein, fibrinogen, and interleukins are potential mediators (or markers) of the eGFR and VTE association.^22,32-34^ Meanwhile, antithrombin may decrease due to its increased urinary loss accompanied with high UACR.^35^ Hypoalbuminemia can increase thromboxane A2 availability which enhances platelet activation and aggregation, consequently facilitating thrombus formation.^36^ All in all, the kidney might trigger the onset of VTE via these biological pathways, particularly in individuals with genetic susceptibility to the disease.

### Clinical implications

CKD affects 9.1% of the general population worldwide.^37^ Our finding underscores the importance of evaluating kidney health in primary practice in VTE and PE prevention, something that is currently not taken place.^1,17,38,39^ Individuals with high genetic risk, such as thrombophilic family members, might be the most relevant population to implement a personalized program of CKD prevention, screening, and management.

### Strengths and limitations

As far as we know, this is the first study to assess the joint effect of genetic background and CKD on VTE risk. The strengths of our study included prospective design, sufficient power, externally validated PRS, deep phenotyping, as well as the robustness of finding across various pre-specified sensitivity analyses. There are several limitations. First, the observational nature of our study cannot establish causality. Second, the UK Biobank participants were mainly healthy and mid-age volunteers and we only included White of European ancestry for genetic data consideration; thus, the findings might not be generalized to other ethnic and age groups. Third, as most of other studies of general populations, eGFR and UACR were obtained once at baseline that may led to misclassification. This would however lead to bias toward the null hypothesis. Further, the CKD-EPI eGFR_cr-cys_ refit equation we used to calculate eGFR tends to be more reliable than the equations contained serum creatinine or cystatin C alone which also frequently used in other observational studies.^40^ Fourth, some VTE cases could have been missed if the event did not occur in the hospital or was not recognized by clinicians, but this ought to be less likely for severe cases, eg, PE.

## Data Availability

The UK Biobank resource is available to bona fide researchers for health-related research in the public interest. All researchers who wish to access the research resource must register with UK Biobank by completing the registration form in the Access Management System

https://bbams.ndph.ox.ac.uk/ams/

## Conclusions

In conclusion, this large prospective cohort study show that eGFR, UACR and more severe KDIGO risk categories independently associated with future risk of VTE and PE. We also demonstrate that even subtle changes in kidney measures can had additive effects across genetically predisposed individuals. Prevention and management of CKD especially for individuals with high genetic risk may reduce the global burden of VTE.

## ACKNOWLEDGMENTS

This study was conducted using the UK Biobank Resource under application number 73684. We appreciate the efforts of each participant and investigator involved in the UK Biobank, as well as the funders that supported this study.

## AUTHOR CONTRIBUTIONS

JZ and XH conceptualized the study; YS and BL were responsible for data curation; YS and BL were responsible for formal analysis; JZ, YS, and XH were responsible for methodology; CS and ZX were responsible for project administration; YS and XH were responsible for resources; YS, ZX and XH were responsible for funding acquisition; YS and XH provided supervision; JZ and XH wrote the original draft; JJC and ZX provided critical feedback on interpretation; all authors reviewed and edited the manuscript.

## DATA AVAILABILITY STATEMENT

The UK Biobank resource is available to bona fide researchers for health-related research in the public interest. All researchers who wish to access the research resource must register with UK Biobank by completing the registration form in the Access Management System (AMS – https://bbams.ndph.ox.ac.uk/ams/).

